# Interleukin- 10 (IL-10) gene polymorphisms and prostate cancer susceptibility: evidence from a meta-analysis

**DOI:** 10.1101/2020.11.09.20228825

**Authors:** Upendra Yadav, Pradeep Kumar, Vandana Rai

## Abstract

Prostate cancer is the second most frequent cancer in men. The frequency of the prostate cancer is greatly varies in different populations of the world. Three common polymorphisms in promoter region of interleukin-10 (IL-10) gene *viz*. -1082 A>G, -819 C>T and -592 C>A are extensively studied in prostate cancer with inconclusive results. So, a meta-analysis was performed to assess the association between these three IL-10 gene polymorphisms and risk of prostate cancer susceptibility. Suitable studies were retrieved by electronic databases search and odds ratios (ORs) with 95% confidence intervals (CIs) were used as association measure. All the statistical analyses were conducted in the Open Meta-Analyst program.

In our meta-analysis we included 17 studies (10,718 samples), 11 studies (8,391 samples) and 13 studies (7,801 samples) for -1082 A>G, -819 C>T and -592 C>A polymorphisms respectively. The result of the -592 C>A polymorphism revealed low heterogeneity with no association in the overall analysis (OR_Avs.C_= 1.05, 95% CI= 0.99-1.12, p= 0.09, I^2^= 35.89%). In ethnicity based stratified analyses, significant association was found in Caucasian population with prostate cancer using allele contrast model (OR_Avs.C_= 1.08, 95% CI= 1.01-1.16, p= 0.02, I^2^= 19.37%), homozygote model (OR_AAvs.CC_= 1.24, 95% CI= 1.00-1.52, p= 0.04, I^2^= 0%), and dominant model (OR_AA+CAvs.CC_= 1.10, 95% CI= 1.00-1.22, p= 0.05, I^2^= 32.57%). No such results were found in the Asian population. In the other two polymorphisms i.e. -1082 A>G and -819 C>T, no significant association with prostate cancer was observed.

In conclusion, results of present meta-analysis suggested that IL-10 -592 C>A polymorphism plays a role in the progression of the prostate cancer in the Caucasian population.

## Introduction

Prostate cancer is the second most common cancer and fifth leading cause of deaths in males worldwide. The worldwide incidence and mortality rate of prostate cancer is 13.5% and 6.7% respectively. The incidence of prostate cancer greatly varies in different regions of the world (highest in Australia/New Zealand and lowest in South Central Asia). While the mortality is highest in Southern Africa and lowest in South Central Asia [1]. The progression of prostate cancer is often slow and it remains localized firstly and later when the cells of the prostatic tissue abnormally proliferate it may spread to nearby tissues and organs and then metastasizes. The etiology of the prostate is least understood despite being a common cancer. A few genetic factors were identified which are consider as the risk for the prostate cancer including age, family history of prostate cancer and ethnicity. Smoking, diet, androgen, and obesity are the secondary factors that may contribute to the risk of prostate cancer [2].

Recently inflammation has been added to the list of secondary factors as it contributes to proliferation, malignancy, angiogenesis, metastasis, modulation of adaptive immunity, and unresponsiveness to hormones and chemotherapeutic agents [3]. Interleukin-10 (IL-10) is an anti-inflammatory cytokine primarily produced by monocytes and to a lesser amount by lymphocytes like-T_H_2, CD4^+^ and CD8^+^ T cells, macrophages, and dendritic cells [4]. IL-10 down regulates the cytokine production of the T_H_1 cells [5]. It enhances the proliferation of B cells, thymocytes, and mast cells and also stimulates the antibody production [6, 7].

The IL-10 cytokine in humans is coded by interlukin-10 (*IL-10*) gene, which is located on the chromosome 1 (1q31-32) and contains 5 exons [8]. Numerous polymorphisms are reported on the *IL-10* gene but clinically important three polymorphisms on the promoter region *viz*. - 1082A>G (rs1800896), -819C>T (rs1800871) and -592C>A (rs1800872) have been extensively studied in different types of cancers like-breast [9], lung [10], gastric [11], cervical [12, 13], and bladder cancer [14].

IL-10 may possibly increase tumor development, as it suppresses the anti-tumor immune response. Involvement of *IL-10* polymorphisms in relation to cancer development is controversial as it has both immune-suppressive as well as anti-angiogenic properties [15]. The role of IL-10 in prostate cancer susceptibility and tumor development is still not clear [16, 17]. A number of association studies were published which evaluated the role of *IL-10* gene polymorphisms in prostate cancer susceptibility [3, 18-23]. These studies investigated the role of *IL-10* gene polymorphisms in the prostate cancer risk with contradicted results. The reason might be due to the limited number of samples or due to the other factors like ethnicities. Therefore, authors performed a meta-analysis to check the association of these three *IL-10* gene polymorphisms with the prostate cancer susceptibility.

## Materials and methods

### Study selection

For the present study, we searched PubMed, Google scholar, and Springer Link databases up to July, 2020. The following keyword were used for the literature search “prostate cancer” along with the combination of “IL-10”, “Interleukin 10”, “IL-10 -1082”, “IL-10 -819”, “IL-10 - 592”. Retrieved articles were searched manually for the additional citations.

### Inclusion and exclusion criteria

The inclusion criteria are-i) case-control study; ii) provided number of alleles and/or genotypes in both case and controls. We excluded the studies which are i) only case reports; ii) not providing enough data to calculate OR and 95%CI; iii) conducted on the animal model; iv) containing overlapping data; and v) review article, editorial, letter to editor etc.

### Date extraction

From all the included articles the following information were retrieved-i) first author’s name; ii) year of publication; iii) ethnicity; iv) country of study; v) number of cases and controls; and vi) number of alleles and genotypes in both cases and controls. All this information were extracted by the authors individually and if any discrepancy were found it was resolved by the discussion.

### Meta-analysis

The odds ratio (OR) with 95% confidence interval (CI) were calculated for the five genetic models *viz*. allele contrast model, dominant model, homozygote model, heterozygote model and recessive model to check the association between prostate cancer risk and *IL-10* gene polymorphisms. The between study heterogeneity was checked by the Q-statistics (p< 0.05 was considered as statistically significant heterogeneity) and I^2^ statistic was used to quantify the inconsistency between study estimates. When a significant heterogeneity was found random effect model [24] was used otherwise fixed effect model [25] was applied. Further analyses were performed on the basis of sub-group analysis based on ethnicity. The evaluation of the symmetry of the funnel plot was used for the identification of the publication bias. Egger’s linear regression method was used for further evaluation of the publication bias [26]. All the statistical analyses were performed in the Open Meta-Analyst program [27]. All p-values were two-tailed with a significance level of 0.05.

## Results

For the present meta-analysis, we followed the PRISMA (Preferred Reporting Items for Systematic Reviews and Meta-Analyses) guideline [28]. Flow chart of article selection was shown in **Fig. 1** with specific reasons. Eighteen studies were found to be eligible for inclusion in the present meta-analysis after applying the inclusion and exclusion criteria [3, 18-23, 29-39].

**Figure 1.**
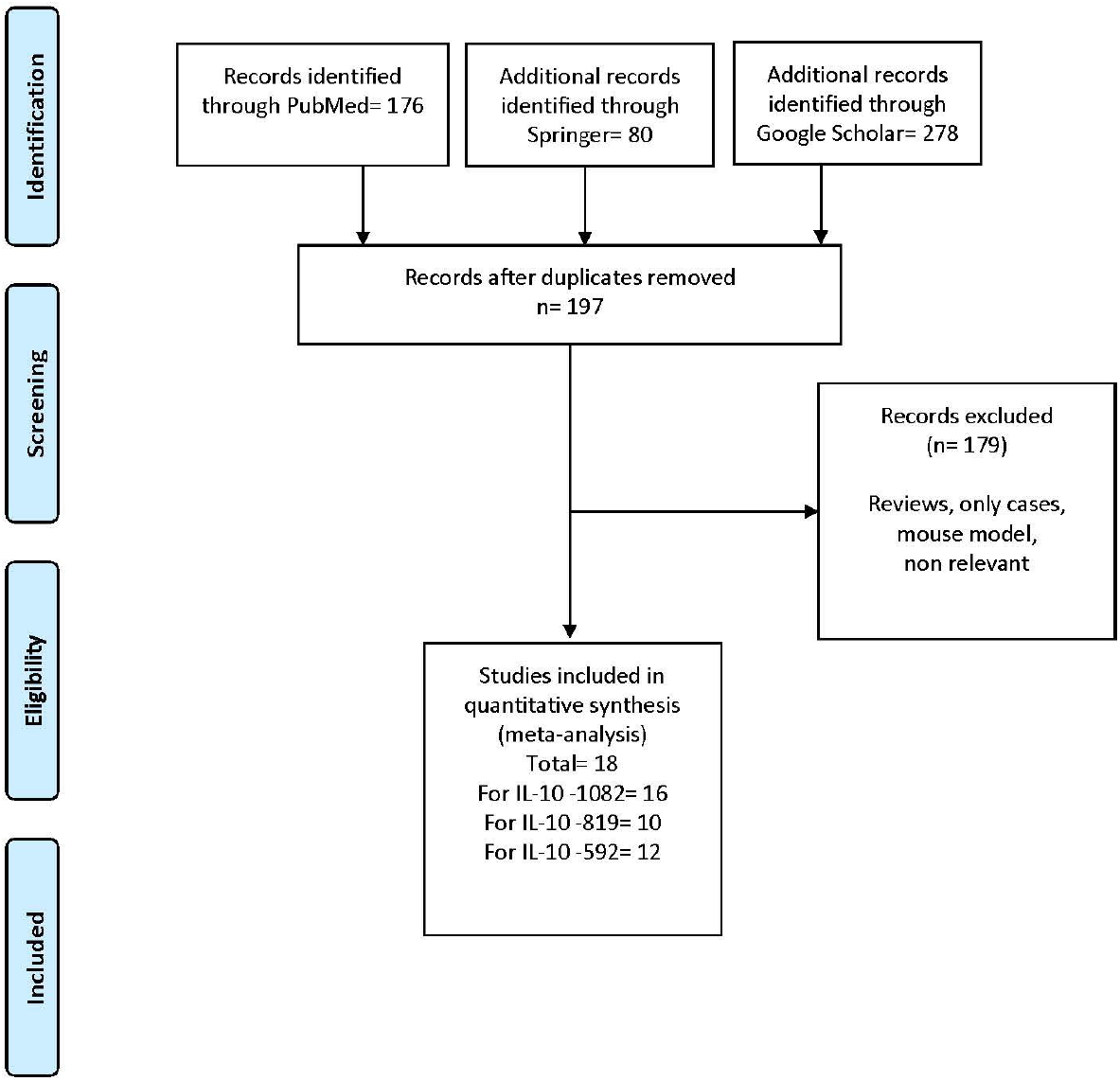
PRISMA Flow diagram of study search and selection process.

### Eligible studies

#### *For IL-10* -1082

A total of 16 studies were included in *IL-10* -1082 meta-analysis [3, 18, 20, 22, 23, 29-39]. The study conducted by Zabaleta et al. (2008)^20^ analyzed two different populations, so we considered them as separate studies. Hence, total 17 studies with 4,937 cases and 5,781 controls were included in this meta-analysis. In the study of Xu et al. [18] only the number of alleles were given. Out of 17 studies, subjects of four studies were Asian, subjects of 11 studies were Caucasian, subjects of one study was Afro-American and another one study subjects was of mixed ethnicity.

#### *For IL-10* -819

Total 10 studies were retrieved and included in *IL-10* -819 meta-analysis (20-23, 29, 30, 32, 34-36). The study conducted by Zabaleta et al. [20] contained two different populations so, we considered them as separate studies. Hence, we have total 11 studies with 3,743 cases and 4,648 controls. Out of selected 11 selected studies, four studies were Asian, five from Caucasian, one Afro-American and one of mixed ethnicity.

#### *For IL-10* -592

Twelve studies were found eligible for *IL-10* -592 meta-analysis [18-23, 30, 33-37]. The study conducted by Zabaleta et al. [20] contained two different populations so we considered them as separate studies. Total 13 studies with 3,771 cases and 4,030 controls were included in the meta-analysis. In the study of Xu et al. [18] only the number of alleles were provided. Out of selected 13 studies, three were Asian, nine from Caucasian, and one belongs to Afro-American ethnicity.

### Meta-analysis

#### *For IL-10* -1082

High heterogeneity was found in allele contrast model with no statistically significant association with prostate cancer (OR_Gvs.A_= 0.98, 95% CI= 0.87-1.10, p= 0.76, I^2^= 75.48%) **(Fig. 2, Table 1)**. Insignificant association were found in all other four genetic models- for dominant model (GG+AG vs. AA) OR= 0.99, 95% CI= 0.81-1.20, p= 0.94, I^2^= 73.54%; for homozygote model (GG vs. AA) OR= 0.93, 95%CI= 0.72-1.22, p= 0.63, I^2^= 73.49; for co-dominant model (AG vs. AA) OR= 1.01, 95%CI= 0.84-1.21, p= 0.89, I^2^= 65.15%; and for recessive model (AA+AG vs. GG) OR= 1.05, 95%CI= 0.86-1.28, p= 0.62, I^2^= 66.71. High heterogeneity was found in all the studies so random effect model was applied **(Table 1)**.

**Table 1.**
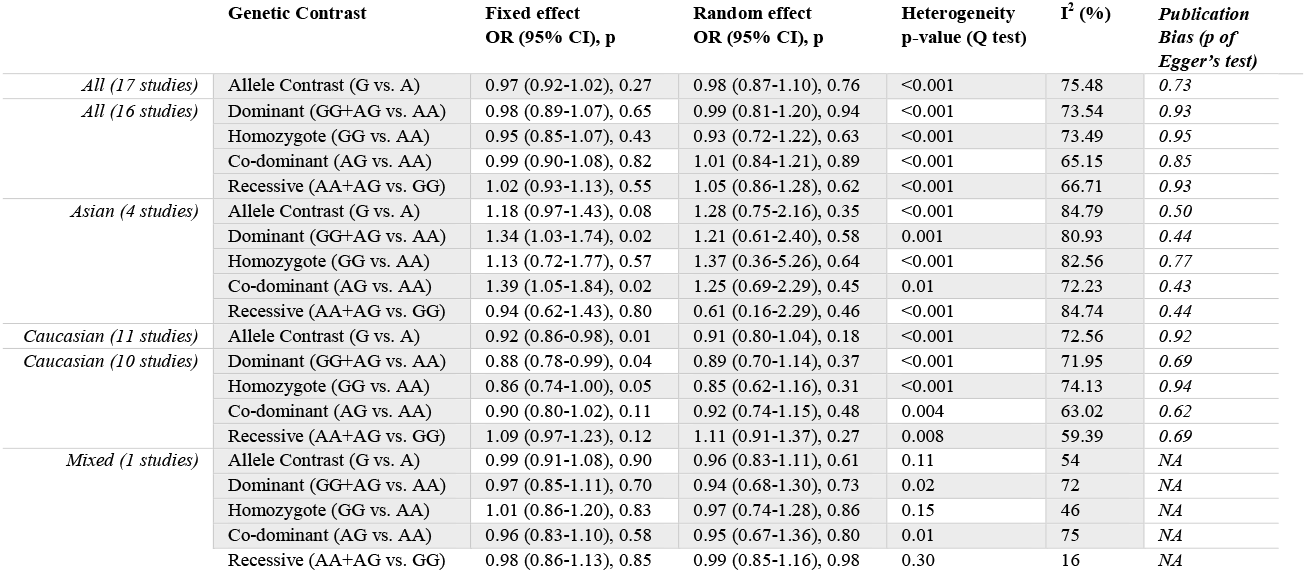
Summary estimates for the odds ratio (OR) of *IL-10* -1082 A>G in various allele/genotype contrasts, the significance level (p value) of heterogeneity test (Q test), I^**2**^ metric and the publication bias.

**Figure 2.**
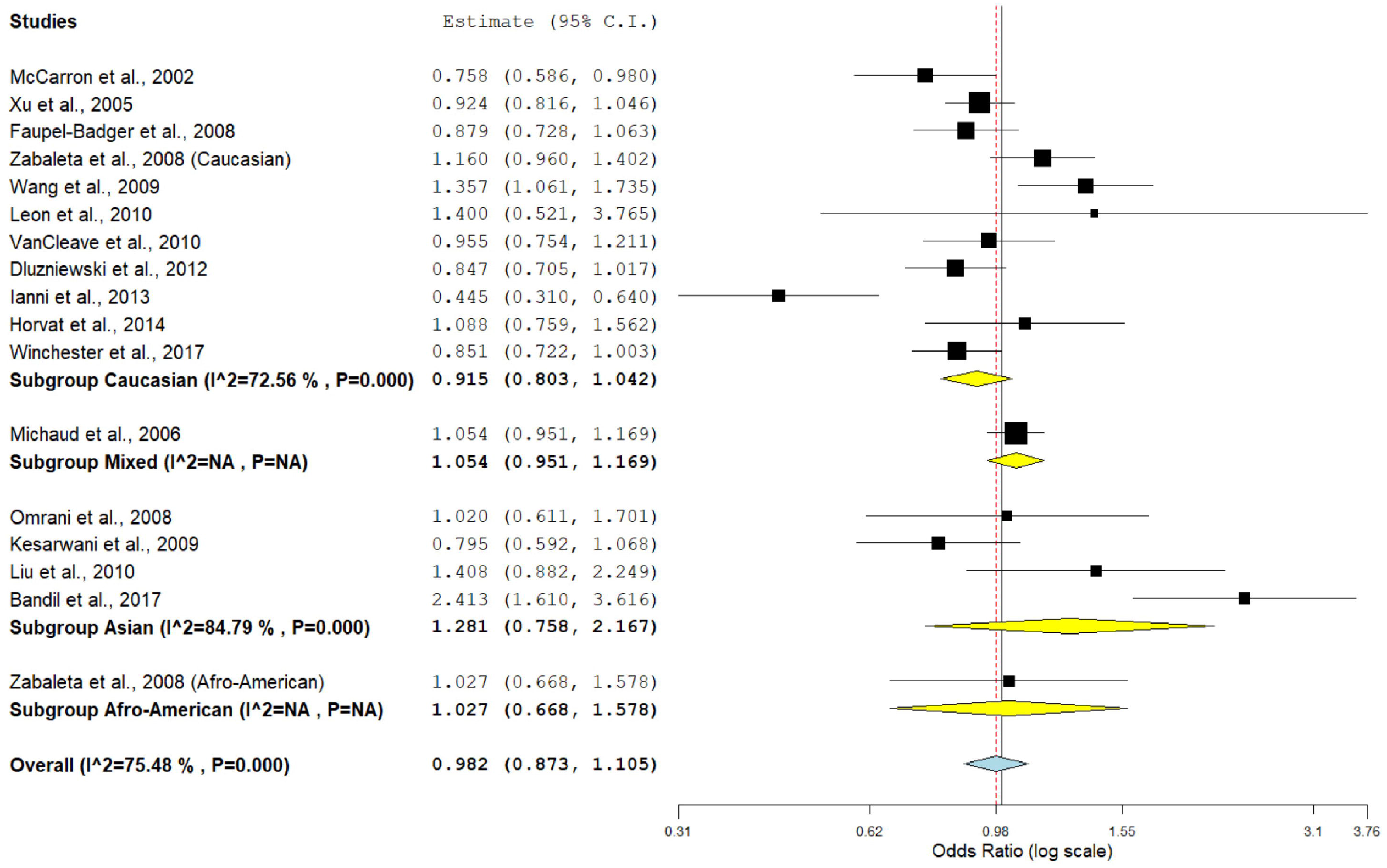
Random effect forest plot of allele contrast model of IL-10 -1082 A>G polymorphism. Results of individual and summary OR estimates, and 95% CI of each study were shown. Horizontal lines represented 95% CI, and dotted vertical lines represent the value of the summary OR.

In subgroup analysis, no significant association was found in any ethnic population using five genetic models and heterogeneity was high in all genetic models **(Table 1)**.

#### *For IL-10* -819

Insignificant association with higher heterogeneity was found in the allele contrast model (OR_Tvs.C_= 1.00, 95%CI= 0.89-1.12, p= 0.98; I^2^= 53.51%) **(Fig. 3, Table 2)**. Similarly, no significant association was observed in any genetic model- for dominant model (TT+CT vs. CC: OR= 1.00, 95%CI= 0.83-1.20, p= 0.96, I^2^= 65.37%), for homozygote model (TT vs. CC: OR= 0.92, 95%CI= 0.79-1.08, p= 0.36, I^2^= 10.71), for co-dominant model (CT vs. CC: OR= 1.01, 95%CI= 0.82-1.23, p= 0.91, I^2^= 66.8%), and for recessive model (CC+CT vs. TT: OR= 1.03, 95%CI= 0.90-1.19, p= 0.60, I^2^= 0%). In the sub-group meta-analyses, no significant association was found in any genetic model in any sub-group **(Table 2)**.

**Table 2.** Summary estimates for the odds ratio (OR) of *IL-10* -819 C>T in various allele/genotype contrasts, the significance level (p value) of heterogeneity test (Q test), I^2^ metric and the publication bias.

| | Genetic Contrast | Fixed effect<br>OR (95% CI), p | Random effect<br>OR (95% CI), p | Heterogeneity<br>p-value (Q test) | $I^2$ (%) | Publication<br>Bias (p of<br>Egger's test) |
| --- | --- | --- | --- | --- | --- | --- |
| *All*<br>*(11 studies)* | Allele Contrast (T vs. C) | 0.98 (0.91-1.05), 0.61 | 1.00 (0.89-1.12), 0.98 | 0.01 | 53.51 | 0.55 |
|  | Dominant (TT+CT vs. CC) | 0.98 (0.89-1.08), 0.75 | 1.00 (0.83-1.20), 0.96 | 0.001 | 65.37 | 0.71 |
|  | Homozygote (TT vs. CC) | 0.92 (0.79-1.08), 0.36 | 0.93 (0.78-1.11), 0.46 | 0.34 | 10.71 | 0.48 |
|  | Co-dominant (CT vs. CC) | 0.99 (0.89-1.09), 0.84 | 1.01 (0.82-1.23), 0.91 | <0.001 | 66.8 | 0.69 |
|  | Recessive (CC+CT vs. TT) | 1.03 (0.90-1.19), 0.60 | 1.03 (0.90-1.19), 0.59 | 0.51 | 0 | 0.79 |
| *Asian*<br>*(4 studies)* | Allele Contrast (T vs. C) | 0.96 (0.84-1.10), 0.62 | 0.97 (0.81-1.17), 0.78 | 0.16 | 41.45 | 0.46 |
|  | Dominant (TT+CT vs. CC) | 0.94 (0.74-1.18), 0.61 | 0.99 (0.60-1.63), 0.99 | 0.005 | 76.73 | 0.35 |
|  | Homozygote (TT vs. CC) | 0.88 (0.67-1.16), 0.38 | 0.88 (0.67-1.16), 0.38 | 0.48 | 0 | 0.63 |
|  | Co-dominant (CT vs. CC) | 0.95 (0.74-1.21), 0.68 | 1.03 (0.56-1.88), 0.91 | <0.001 | 82.07 | 0.26 |
|  | Recessive (CC+CT vs. TT) | 1.03 (0.83-1.26), 0.76 | 1.03 (0.83-1.26), 0.77 | 0.44 | 0 | 0.43 |
| *Caucasian*<br>*(5 studies)* | Allele Contrast (T vs. C) | 1.09 (0.97-1.22), 0.11 | 1.09 (0.90-1.32), 0.35 | 0.04 | 58 | 0.98 |
|  | Dominant (TT+CT vs. CC) | 1.11 (0.96-1.28), 0.13 | 1.10 (0.86-1.41), 0.42 | 0.03 | 60.76 | 0.94 |
|  | Homozygote (TT vs. CC) | 1.16 (0.88-1.53), 0.28 | 1.16 (0.86-1.55), 0.32 | 0.36 | 7.22 | 0.83 |
|  | Co-dominant (CT vs. CC) | 1.09 (0.94-1.27), 0.21 | 1.09 (0.84-1.40), 0.49 | 0.04 | 58.77 | 0.97 |
|  | Recessive (CC+CT vs. TT) | 0.88 (0.68-1.15), 0.38 | 0.89 (0.68-1.16), 0.41 | 0.47 | 0 | 0.81 |
| *Mixed*<br>*(1 studies)* | Allele Contrast (T vs. C) | 0.90 (0.81-1.01), 0.07 | 0.90 (0.81-1.01), 0.07 | 0.73 | 0 | NA |
|  | Dominant (TT+CT vs. CC) | 0.90 (0.79-1.03), 0.14 | 0.90 (0.79-1.03), 0.14 | 0.71 | 0 | NA |
|  | Homozygote (TT vs. CC) | 0.82 (0.64-1.05), 0.12 | 0.82 (0.64-1.05), 0.12 | 0.76 | 0 | NA |
|  | Co-dominant (CT vs. CC) | 0.92 (0.80-1.06), 0.28 | 0.92 (0.80-1.06), 0.28 | 0.68 | 0 | NA |
|  | Recessive (CC+CT vs. TT) | 1.18 (0.93-1.49), 0.17 | 1.18 (0.93-1.49), 0.16 | 0.83 | 0 | NA |

**Figure 3.**
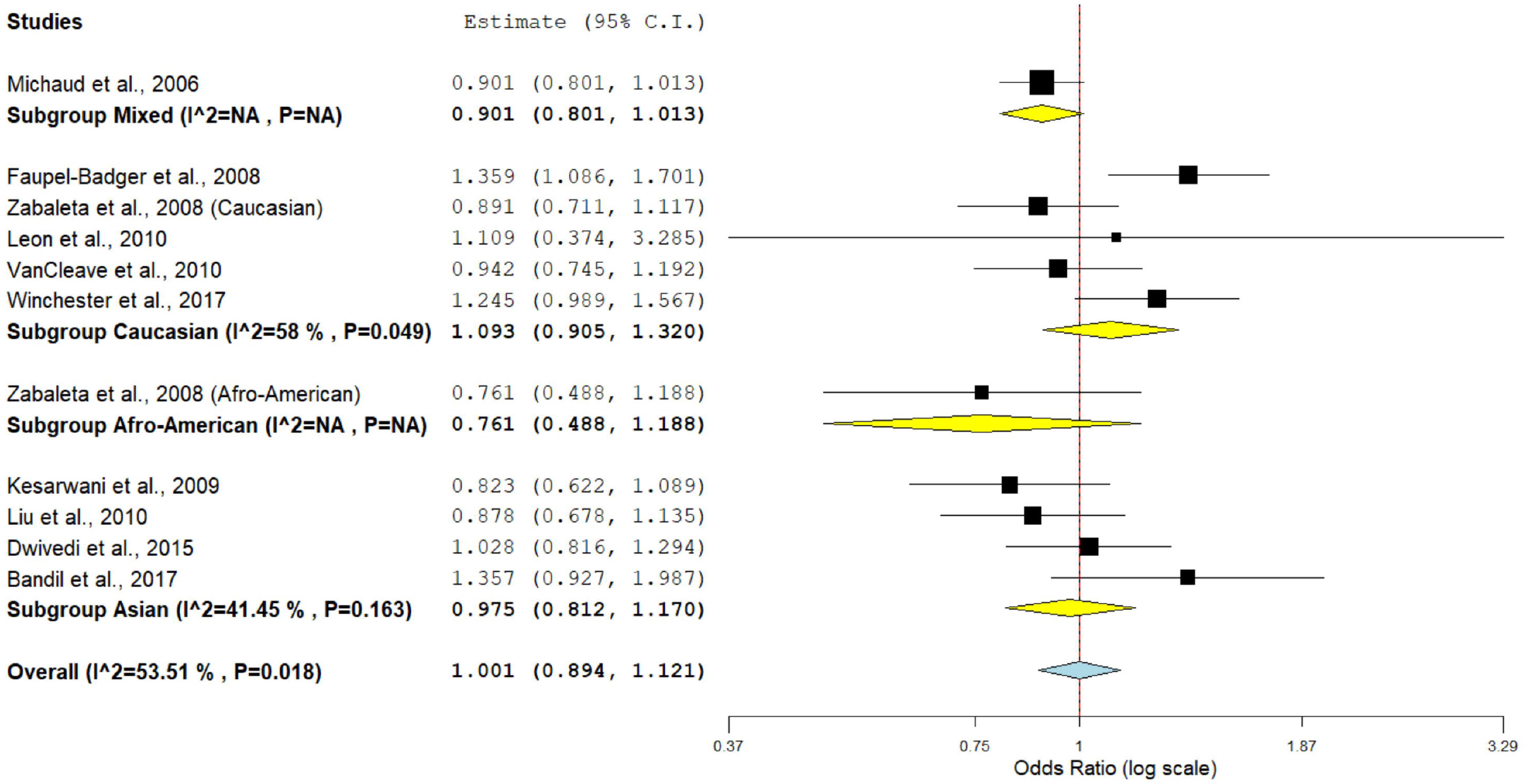
Random effect forest plot of allele contrast model of IL-10 -819 C>T polymorphism.

#### *For IL-10* -592

Non-significant association was found in the allele contrast model with low heterogeneity (OR_Avs.C_= 1.05, 95%CI= 0.99-1.12, p= 0.09, I^2^= 35.89) **(Fig. 4, Table 3)**. Similarly no significant association with low heterogeneity was observed in all genetic models- for dominant model (AA+CA vs. CC: OR= 1.07, 95%CI= 0.97-1.17, p= 0.14, I^2^= 23.57%), for homozygote model (AA vs. CC: OR= 1.10, 95%CI= 0.93-1.30, p= 0.26, I^2^= 30.19%), for co-dominant model (CA vs. CC: OR= 1.05, 95%CI= 0.96-1.16, p= 0.24, I^2^= 22.36%) and for recessive model (CC+CA vs. AA: OR= 0.93, 95%CI= 0.79-1.09, p= 0.37, I^2^= 37.04%) **(Table 3)**.

**Table 3.** Summary estimates for the odds ratio (OR) of *IL-10* -592 C>A in various allele/genotype contrasts, the significance level (p value) of heterogeneity test (Q test), I^**2**^ metric and the publication bias.

| | Genetic Contrast | Fixed effect<br>OR (95% CI), p | Random effect<br>OR (95% CI), p | Heterogeneity<br>p-value (Q test) | $I^2$ (%) | Publication<br>Bias (p of<br>Egger's test) |
| --- | --- | --- | --- | --- | --- | --- |
| All (13 studies) | Allele Contrast (A vs. C) | 1.05 (0.99-1.12), 0.09 | 1.05 (0.96-1.14), 0.26 | 0.09 | 35.89 | 0.39 |
| All (12 studies) | Dominant (AA+CA vs. CC) | 1.07 (0.97-1.17), 0.14 | 1.06 (0.95-1.18), 0.26 | 0.21 | 23.57 | 0.38 |
| All (12 studies) | Homozygote (AA vs. CC) | 1.10 (0.93-1.30), 0.26 | 1.09 (0.88-1.36), 0.39 | 0.15 | 30.19 | 0.72 |
| All (12 studies) | Co-dominant (CA vs. CC) | 1.05 (0.96-1.16), 0.24 | 1.05 (0.94-1.18), 0.34 | 0.22 | 22.36 | 0.74 |
| All (12 studies) | Recessive (CC+CA vs. AA) | 0.93 (0.79-1.09), 0.37 | 0.93 (0.74-1.15), 0.51 | 0.09 | 37.04 | 0.79 |
| Asian (3 studies) | Allele Contrast (A vs. C) | 0.94 (0.80-1.10), 0.49 | 0.91 (0.70-1.19), 0.52 | 0.07 | 61.59 | 0.47 |
|  | Dominant (AA+CA vs. CC) | 0.96 (0.77-1.20), 0.74 | 0.96 (0.77-1.20), 0.74 | 0.45 | 0 | 0.91 |
|  | Homozygote (AA vs. CC) | 0.87 (0.63-1.20), 0.42 | 0.80 (0.42-1.52), 0.51 | 0.03 | 71.33 | 0.49 |
|  | Co-dominant (CA vs. CC) | 0.99 (0.78-1.26), 0.98 | 1.01 (0.76-1.34), 0.93 | 0.24 | 28.8 | 0.46 |
|  | Recessive (CC+CA vs. AA) | 1.13 (0.84-1.51), 0.40 | 1.28 (0.62-2.63), 0.49 | 0.006 | 80.53 | 0.42 |
| Caucasian (9 studies) | Allele Contrast (A vs. C) | **1.08 (1.01-1.16), 0.02** | 1.09 (1.00-1.18), 0.03 | 0.27 | 19.37 | 0.79 |
| Caucasian (8 studies) | Dominant (AA+CA vs. CC) | **1.10 (1.00-1.22), 0.05** | 1.10 (0.97-1.25), 0.13 | 0.16 | 32.57 | 0.91 |
| Caucasian (8 studies) | Homozygote (AA vs. CC) | **1.24 (1.00-1.52), 0.04** | 1.23 (1.00-1.52), 0.04 | 0.65 | 0 | 0.54 |
| Caucasian (8 studies) | Co-dominant (CA vs. CC) | 1.08 (0.97-1.20), 0.15 | 1.08 (0.94-1.23), 0.25 | 0.18 | 30.32 | 0.90 |
| Caucasian (8 studies) | Recessive (CC+CA vs. AA) | **0.83 (0.68-1.02), 0.07** | **0.83 (0.68-1.02), 0.08** | **0.71** | **0** | **0.42** |

**Figure 4.**
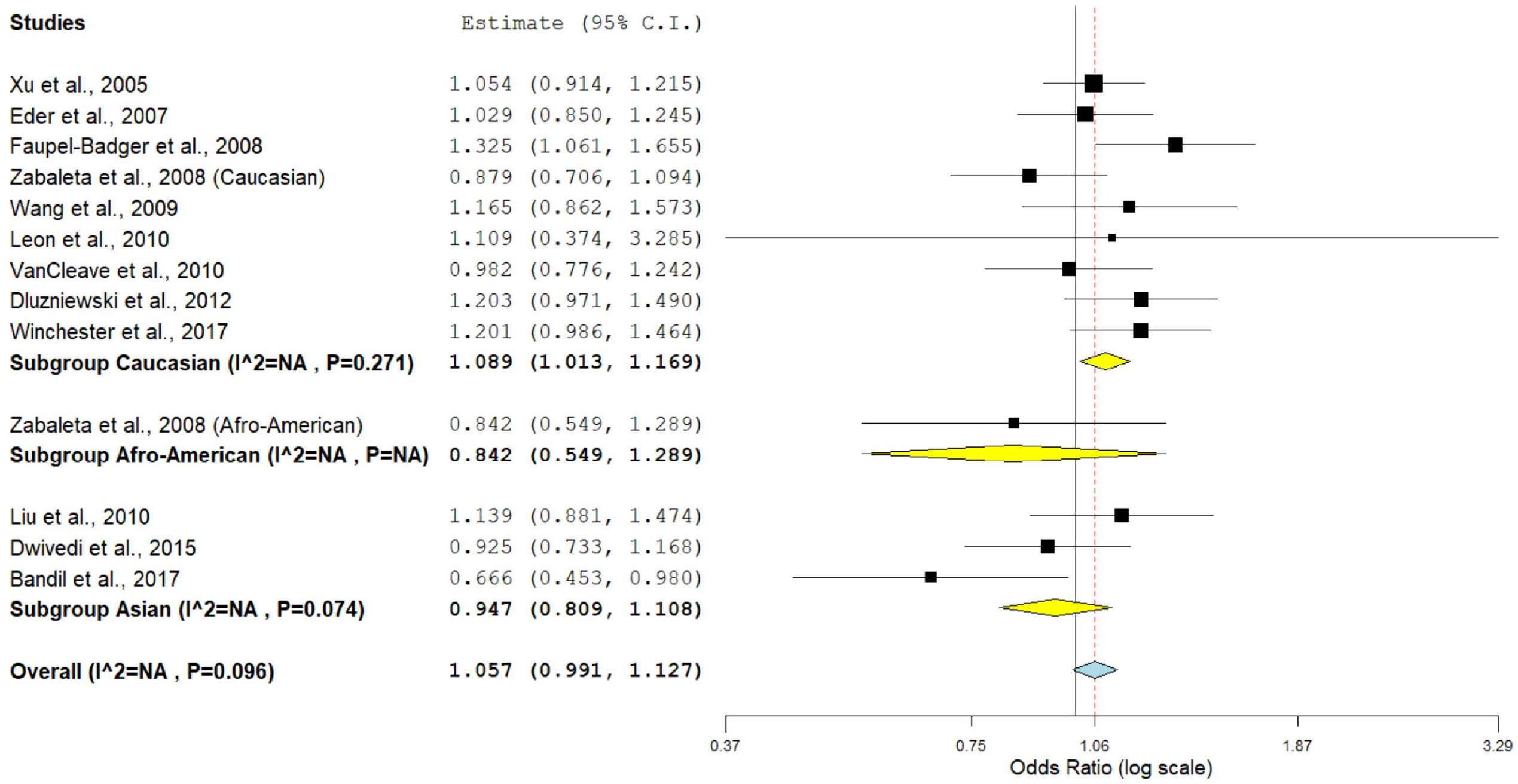
Random effect forest plot of allele contrast model of IL-10 -592 C>A polymorphism.

Ethnicity based sub-group analyses were performed and no statistically significant association was found in any genetic model in the Asian sub-group **(Table 3)**. However, in the Caucasian sub-group meta-analysis, marginal association was observed in the allele contrast model (OR_Avs.C_= 1.08, 95%CI= 1.01-1.16, p= 0.02, I^2^= 19.37%). Similar association was also found in the dominant model (AA+CA vs. CC) OR= 1.10, 95%CI= 1.00-1.22, p= 0.05, I^2^= 32.57%; and homozygote model (AA vs. CC) OR= 1.24, 95%CI= 1.00-1.52, p= 0.04, I^2^= 0% **(Fig. 5, Table 3)**. But no association was observed in the other two models i.e. co-dominant (CA vs. CC) and recessive (CC+CA vs. AA) models **(Table 3)**.

**Figure 5.**
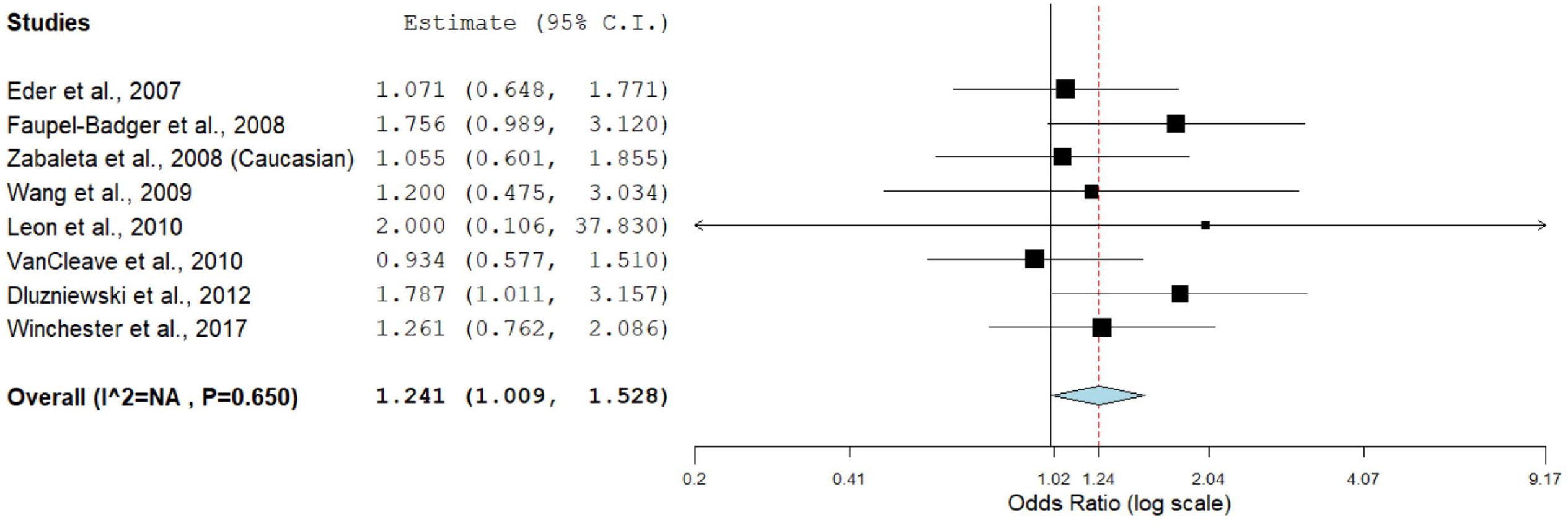
Fixed effect forest plot of homozygote model of IL-10 -592 C>A polymorphism in the Caucasian population.

### Sensitivity analysis

The sensitivity analysis was performed by the removal of the studies in which the control population were not in the Hardy-Weinberg equilibrium (HWE). For *IL-10* -1082 four studies were deviated from the HWE [31, 20 (Afro-American), 32, 35]. Removal of these studies has no effect on the overall meta-analysis or subgroup analyses. For *IL-10* -819 two studies were not in HWE [22, 36]. When we performed meta-analysis after removal of these studies no effect was observed in overall meta-analysis or sub-group meta-analyses. For *IL-10* -592 only one study [22] was deviated from HWE. When this study was removed from the analysis, we found that the heterogeneity was decreased and a marginally significant association was observed in the allele contrast model (OR_Avs.C_= 1.07, 95%CI= 1.00-1.14, p= 0.03, I^2^= 19.37%). While no such effect was found in any other genetic model in overall as well as in the sub-group meta-analyses.

### Publication bias

Publication bias was assessed by the evaluation of the funnel plot and a symmetry in the funnel plot demonstrates lack of publication bias. Symmetrical funnel plots were found in all genetic models (**Fig. 6**). To confirm this statistically, we performed the Egger’s test and the p-value of Egger’s test is greater than 0.05 in all the genetic models in all the selected polymorphisms both in overall as well as in sub-group analyses (**Table 1-3**).

**Figure 6.**
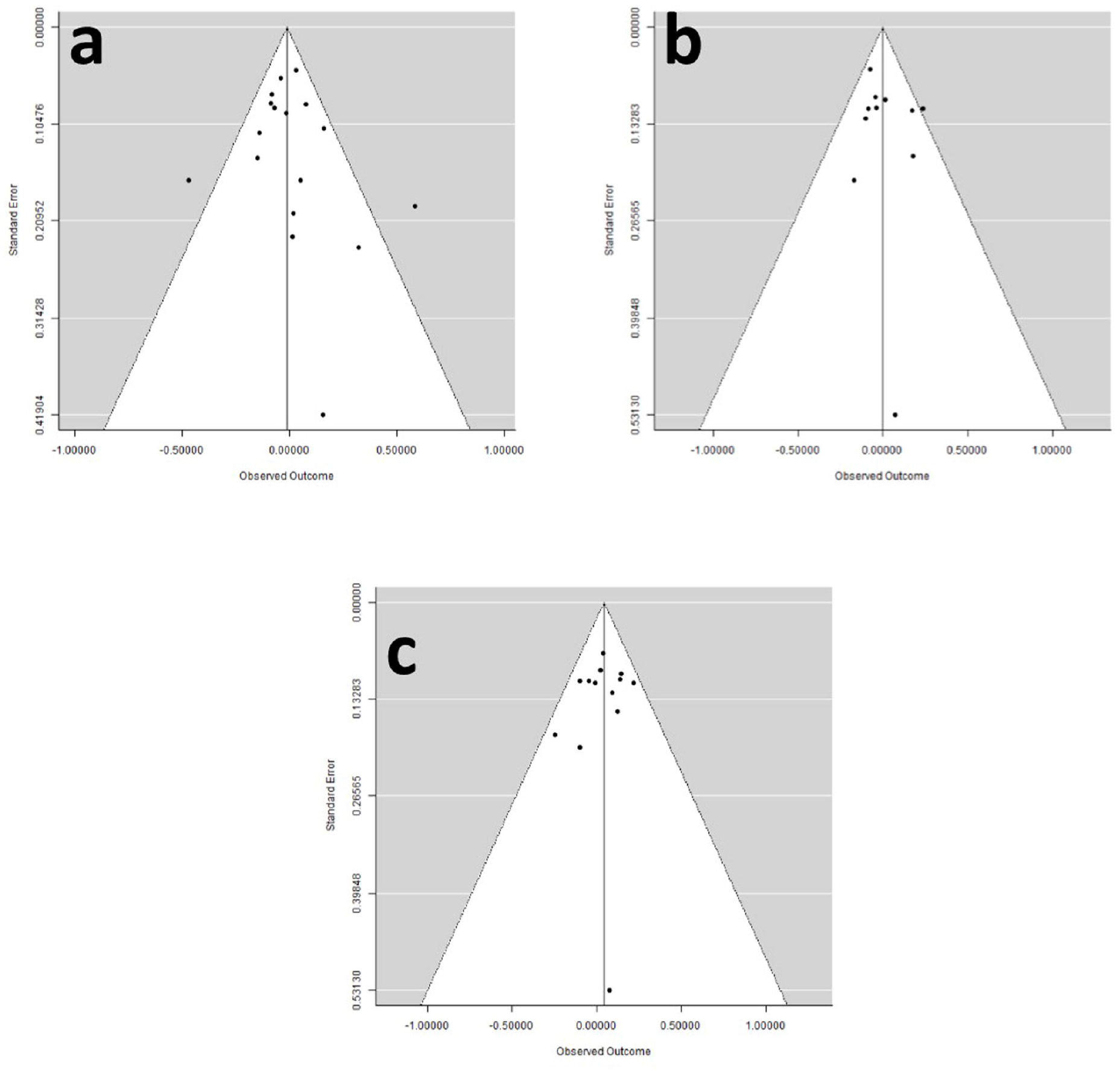
Funnel plots of standard error by log odds ratio for a) IL-10-1082 A>G; b) IL-10 -819 C>T; and c) IL-10 -592 C>A.

## Discussion

In the present study, we examined the role of three common promoter polymorphisms (−1082A>G, -819C>T and -592C>A) of the *IL-10* gene in the susceptibility of the prostate cancer. The result of the present study indicates that the -592 polymorphism of the *IL-10* gene shows significant association with the prostate cancer risk in the Caucasian population in allele contrast (OR_Avs.C_= 1.08, 95%CI= 1.01-1.16, p= 0.02, I^2^= 19.37%), dominant model (OR_AA+CAvs.CC_= 1.10, 95%CI= 1.00-1.22, p= 0.05, I^2^= 32.57%) and homozygote model (OR_AAvs.CC_= 1.24, 95%CI= 1.00-1.52, p= 0.04, I^2^= 0%). No significant association was observed in overall meta-analysis or in the sub-group analyses in the other two polymorphisms.

IL-10 cytokine demonstrates both anti-inflammatory and anti-angiogenic properties. The anti-inflammatory mechanisms of IL-10 are suspected to have pro-tumorigenic potential by allowing cancerous cells to avoid immune surveillance. The IL-10 also have anti-angiogenic properties and it decreases the tumor growth and angiogenesis both in animals and in *in-vitro* models [30]. The expression of the IL-10 cytokine is influenced by the promoter polymorphisms (−1082A>G, -819C>T and -592C>A). Reduced level of IL-10 is reported in the presence of *IL-10* -1082G/-819T/-592A haplotype [19, 40, 41]. Several studies were published examining the role of *IL-10* gene polymorphisms and susceptibility to prostate cancer but the results were inconclusive. The reasons for this is not clear however, several possibilities may exist. The first reason might be due to the differences in the distribution of the *IL-10* gene polymorphisms amongst the different ethnic populations. In addition, the prostate cancer is a multi-factorial disease and the exposure of various environmental factors may influence the expression of some genes e.g. the interaction of promoter genetic variants of *IL-10* gene with exposure to tobacco may affect the severity as well as susceptibility of prostate cancer [42].

During literature search we found five meta-analyses, which examined the role of *IL-10* gene polymorphisms in the susceptibility of prostate cancer [17, 43-46]. Shao et al. [43] have found modest association of -819 C>T (OR_Tvs.C_= 1.16, 95% CI= 1.03-1.30, p= 0.01) and -592 C>A (OR_Avs.C_= 1.13, 95% CI= 1.01-1.26, p= 0.03) polymorphisms with prostate cancer. Three meta-analyses [17, 44, 45], did not report any association between *IL-10* gene polymorphisms and prostate cancer risk. Men et al. [46] included 17 studies in their meta-analysis and reported marginally significant association of -1082A>G (OR=1.10, 95% CI= 1.00-1.21, p= 0.05, I^2^= 64.3%). In the present meta-analysis, we included three polymorphisms (−1082A>G, -819C>T and -592C>A) with 18 studies and found no association between these polymorphisms and prostate cancer. Further analysis were performed on the basis of the ethnicity and we found that the -592C>A polymorphism statistically associated with the risk of prostate cancer in the Caucasian population.

Meta-analysis is an statistical tool for combining the studies with low sample size and low power. In recent years meta-analysis is frequently used to check the association between alleles/genotypes or some environmental factors with some diseases/disorders. In past few years a number of meta-analyses were published like – recurrent pregnancy loss [47], Down syndrome [48, 49], neural tube defects [50], Glucose 6-phosphate dehydrogenase deficiency [51], osteoporosis [52], epilepsy [53], bipolar disorder [54], schizophrenia [55, 56], Alzheimer [57], breast cancer [58-60], colorectal cancer [61], digestive tract cancer [62], esophageal cancer [63], and prostate cancer [64].

Our meta-analysis has few advantages over the previously published meta-analyses such as – i) this is the largest study on the basis of number of included studies (18 studies) and sample size; ii) included three polymorphisms (−1082A>G, -819C>T and -592C>A); iii) absence of publication bias; and iv) included studies from all the ethnicities. At the same time we want to narrate few limitations associated with our meta-analysis like – i) included only studies published in English; ii) only genetic factors were considered; and iii) crude odds ratio was used.

## Conclusions

The result of our meta-analysis showed that the -592C>A polymorphism is significantly associated with the risk of prostate cancer in the Caucasian population, while no such effect is found in other ethnicities or in the other gene polymorphisms. For the future studies we suggest that one should also measure the level of IL-10 expression because these polymorphisms significantly influences the level of IL-10 cytokine. Furthermore, if gene-environmental interaction is considered in the future studies it would be an added advantage to elucidate the clear picture of the *IL-10* gene polymorphisms in the development of the prostate cancer.

## Data Availability

All the data is given in the manuscript.

## Acknowledgements

Upendra Yadav is highly grateful to the VBS Purvanchal University, Jaunpur, for providing financial assistance to him in the form of PDF.

## References

1. Bray F, Ferlay J, Soerjomataram I, Siegel RL, Torre LA, Jemal A. Global cancer statistics 2018: GLOBOCAN estimates of incidence and mortality worldwide for 36 cancers in 185 countries. CA Cancer J Clin 2018;68(6):394–424.

2. Hsing AW, Chokkalingam AP. Prostate cancer epidemiology. Front Biosci 2006;11:1388–413.

3. McCarron SL, Edwards S, Evans PR, Gibbs R, Dearnaley DP, Dowe A, et al. Influence of cytokine gene polymorphisms on the development of prostate cancer. Cancer Res 2002;62:3369–72.

4. Saraiva M, O’Garra A. The regulation of IL-10 production by immune cells. Nat Rev Immunol 2010;10(3):170–81.

5. Mosser DM, Zhang X. Interleukin-10: new perspectives on an old cytokine. Immunol Rev 2008;226:205–18.

6. Moore KW, de Waal Malefyt R, Coffman RL, O’Garra A. Interleukin-10 and the interleukin-10 receptor. Annu Rev Immunol 2001;19:683–765.

7. Sabat R, Grutz G, Warszawska K, Kirsch S, Witte E, Wolk K, et al. Biology of interleukin-10. Cytokine Growth Factor Rev 2010;21(5):331–44.

8. Eskdale J, Kube D, Tesch H, Gallagher G. Mapping of the human IL-10 gene and further characterisation of the 5’ flanking sequence. Immunogenetics 1997;46:120–8.

9. Li M, Yue C, Zuo X, Jin G, Wang G, Guo H, et al. The effect of interleukin 10 polymorphisms on breast cancer susceptibility in Han women in Shaanxi Province. PLoS One 2020;15(5):e0232174.

10. Shih CM, Lee YL, Chiou HL, Hsu WF, Chen WE, Chou MC, et al. The involvement of genetic polymorphism of IL-10 promoter in non-small cell lung cancer. Lung Cancer 2005;50(3):291–7.

11. Kumar S, Kumari N, Mittal RD, Mohindra S, Ghoshal UC. Association between pro-(IL-8) and anti-inflammatory (IL-10) cytokine variants and their serum levels and H. pylori-related gastric carcinogenesis in northern India. Meta Gene 2015;6:9–16.

12. Datta A, Tuz Zahora F, Abdul Aziz M, Sarowar Uddin M, Ferdous M, Shalahuddin Millat M, et al. Association study of IL10 gene polymorphisms (rs1800872 and rs1800896) with cervical cancer in the Bangladeshi women. Int Immunopharmacol 2020;89(Pt B):107091.

13. Pereira APL, Trugilo KP, Okuyama NCM, Sena MM, Couto-Filho JD, Watanabe MAE, et al. IL-10 c.-592C>A (rs1800872) polymorphism is associated with cervical cancer. J Cancer Res Clin Oncol 2020;146(8):1971–8.

14. Ahirwar D, Mandhani A, Mittal RD. Interleukin-10 G-1082A and C-819T polymorphisms as possible molecular markers of urothelial bladder cancer. Arch Med Res 2009;40(2):97–102.

15. Plantinga TS, Costantini I, Heinhuis B, Huijbers A, Semango G, Kusters B, et al. A promoter polymorphism in human interleukin-32 modulates its expression and influences the risk and the outcome of epithelial cell-derived thyroid carcinoma. Carcinogenesis 2013;34:1529–35.

16. Howell WM, Rose-Zerilli MJ. Interleukin-10 polymorphisms, cancer susceptibility and prognosis. Fam Cancer 2006;5:143–9.

17. Zou YF, Wang F, Feng XL, Tian YH, Tao JH, Pan FM, et al. Lack of association of IL-10 gene polymorphisms with prostate cancer: evidence from 11,581 subjects. Eur J Cancer 2011;47(7):1072–9.

18. Xu J, Lowey J, Wiklund F, Sun J, Lindmark F, Hsu FC, et al. The interaction of four genes in the inflammation pathway significantly predicts prostate cancer risk. Cancer Epidemiol Biomarkers Prev 2005;14(11 Pt 1):2563–8.

19. Eder T, Mayer R, Langsenlehner U, Renner W, Krippl P, Wascher TC et al. Interleukin-10 [ATA] promoter haplotype and prostate cancer risk: a population-based study. Eur J Cancer 2007;43:472–5.

20. Zabaleta J, Lin HY, Sierra RA, Hall MC, Clark PE, Sartor OA, et al. Interactions of cytokine gene polymorphisms in prostate cancer risk. Carcinogenesis 2008;29(3):573–8.

21. Dwivedi S, Goel A, Khattri S, Mandhani A, Sharma P, Misra S, et al. Genetic variability at promoters of IL-18 (pro-) and IL-10 (anti-) inflammatory gene affects susceptibility and their circulating serum levels: An explorative study of prostate cancer patients in North Indian populations. Cytokine 2015;74(1):117–22.

22. Bandil K, Singhal P, Dogra A, Rawal SK, Doval DC, Varshney AK, et al. Association of SNPs/haplotypes in promoter of TNF A and IL-10 gene together with life style factors in prostate cancer progression in Indian population. Inflamm Res 2017;66(12):1085–97.

23. Winchester DA, Till C, Goodman PJ, Tangen CM, Santella RM, Johnson-Pais TL, et al. Association between variants in genes involved in the immune response and prostate cancer risk in men randomized to the finasteride arm in the Prostate Cancer Prevention Trial. Prostate 2017;77(8):908–19.

24. DerSimonian R, Laird N. Meta-analysis in clinical trials. Control Clin Trials 1986;7:177–88.

25. Mantel N, Haenszel W. Statistical aspects of the analysis of data from retrospective studies of disease. J Natl Cancer Inst 1959;22(4):719–48.

26. Egger M, Smith DJ, Schneider M, Minder C. Bias in meta-analysis detected by a simple, graphical test. BMJ 1997;315(7109): 629–34.

27. Wallace BC, Dahabreh IJ, Trikalinos TA, Lau J, Trow P, Schmid CH. Closing the gap between methodologists and end-users: R as a computational back-end. J Stat Software 2013;49:1–15.

28. Moher D, Liberati A, Tetzlaff J, Altman DG, The PRISMA Group. Preferred Reporting Items for Systematic Reviews and Meta-Analyses: The PRISMA Statement. PLoS Med 2009;6(6):e1000097.

29. Michaud DS, Daugherty SE, Berndt SI, Platz EA, Yeager M, Crawford ED, et al. Genetic polymorphisms of interleukin-1B (IL-1B), IL-6, IL-8, and IL-10 and risk of prostate cancer. Cancer Res 2006;66(8):4525–30.

30. Faupel-Badger JM, Kidd LC, Albanes D, Virtamo J, Woodson K, Tangrea JA. Association of IL-10 polymorphisms with prostate cancer risk and grade of disease. World J Urol 2008;19:119–24.

31. Omrani MD, Bazargani S, Bageri M. Interlukin-10, Interferon-γ and Tumor Necrosis Factor-α Genes Variation in Prostate Cancer and Benign Prostatic Hyperplasia. Curr Urol 2008;2:175–80.

32. Kesarwani P, Ahirwar DK, Mandhani A, Singh AN, Dalela D, Srivastava AN, et al. IL-10 - 1082 G>A: a risk for prostate cancer but may be protective against progression of prostate cancer in North Indian cohort. World J Urol 2009;27(3):389–96.

33. Wang MH, Helzlsouer KJ, Smith MW, Hoffman-Bolton JA, Clipp SL, Grinberg V, et al. Association of IL10 and other immune response- and obesity-related genes with prostate cancer in CLUE II. Prostate 2009;69(8):874–85.

34. Leon A, Leon V, Garcia J, and Urrutia M. Interleukin gene polymorphism in patients with prostate cancer. Tissue Antigens 2010;75:637.

35. Liu J, Song B, Bai X, Liu W, Li Z, Wang J, et al. Association of genetic polymorphisms in the interleukin-10 promoter with risk of prostate cancer in Chinese. BMC Cancer 2010;10:456.

36. VanCleave TT, Moore JH, Benford ML, Brock GN, Kalbfleisch T, Baumgartner RN, et al. Interaction among variant vascular endothelial growth factor (VEGF) and its receptor in relation to prostate cancer risk. Prostate 2010;70(4):341–52.

37. Dluzniewski PJ, Wang MH, Zheng SL, De Marzo AM, Drake CG, Fedor HL, et al. Variation in IL10 and other genes involved in the immune response and in oxidation and prostate cancer recurrence. Cancer Epidemiol Biomarkers Prev 2012;21(10):1774–82.

38. Ianni M, Porcellini E, Carbone I, Potenzoni M, Pieri AM, Pastizzaro CD, et al. Genetic factors regulating inflammation and DNA methylation associated with prostate cancer. Prostate Cancer Prostatic Dis 2013;16(1):56–61.

39. Horvat V, Mandić S, Marczi S, Mrčela M, Galić J. Association of IL-1β and IL-10 Polymorphisms with Prostate Cancer Risk and Grade of Disease in Eastern Croatian Population. Coll Antropol 2015;39(2):393–400.

40. Turner DM, Williams DM, Sankaran D, Lazarus M, Sinnott PJ, Hutchinson IV. An investigation of polymorphism in the interleukin-10 gene promoter. Eur J Immunogenet 1997;24:1–8.

41. Crawley E, Woo P, Isenberg DA. Single nucleotide polymorphic haplotypes of the interleukin-10 5’ flanking region are not associated with renal disease or serology in Caucasian patients with systemic lupus erythematosus. Arthritis Rheum 1999;42:2017–8.

42. Dwivedi S, Singh S, Goel A, Khattri S, Mandhani A, Sharma P, et al. Pro-(IL-18) and Anti-(IL-10) Inflammatory Promoter Genetic Variants (Intrinsic Factors) with Tobacco Exposure (Extrinsic Factors) May Influence Susceptibility and Severity of Prostate Carcinoma: A Prospective Study. Asian Pac J Cancer Prev 2015;16(8):3173–81.

43. Shao N, Xu B, Mi YY, Hua LX. IL-10 polymorphisms and prostate cancer risk: a meta-analysis. Prostate Cancer Prostatic Dis 2011;14(2):129–35.

44. Yu Z, Liu Q, Huang C, Wu M, Li G. The interleukin 10 -819C/T polymorphism and cancer risk: a HuGE review and meta-analysis of 73 studies including 15,942 cases and 22,336 controls. OMICS 2013;17(4):200–14.

45. Chen H, Tang J, Shen N, Ren K. Interleukin 10 gene rs1800896 polymorphism is associated with the risk of prostate cancer. Oncotarget 2017;8(39):66204–14.

46. Men T, Yu C, Wang D, Liu F, Li J, Qi X, et al. The impact of interleukin-10 (IL-10) gene 4 polymorphisms on peripheral blood IL-10 variation and prostate cancer risk based on published studies. Oncotarget 2017;8(28):45994–6005.

47. Rai V. Methylenetetrahydrofolate reductase gene A1298C polymorphism and susceptibility to recurrent pregnancy loss: a meta-analysis. Cell Mol Biol (Noisy-le-grand) 2014;60(2):27–34.

48. Rai V, Yadav U, Kumar P, Yadav SK, Mishra OP. Maternal methylenetetrahydrofolate reductase C677T polymorphism and down syndrome risk: a meta-analysis from 34 studies. PLoS One 2014;9(9):e108552.

49. Rai V, Kumar P. Fetal MTHFR C677T polymorphism confers no susceptibility to Down Syndrome: evidence from meta-analysis. Egyptian J Med Hum Genet 2018;19:53–8.

50. Yadav U, Kumar P, Yadav SK, Mishra OP, Rai V. “Polymorphisms in folate metabolism genes as maternal risk factor for neural tube defects: an updated meta-analysis”. Metab Brain Dis 2015;30(1):7–24.

51. Kumar P, Yadav U, Rai V. Prevalence of Glucose-6-phosphate dehydrogenase deficiency in India: an updated meta-analysis. Egypt J Med Hum Genet 2016;17:295–302.

52. Yadav U, Kumar P, Rai V. Vitamin D receptor (VDR) gene FokI, BsmI, ApaI and TaqI polymorphisms and osteoporosis risk: a meta-analysis. Egypt J Med Hum Genet 2020 (In press) doi.org/10.1186/s43042-020-00057-5.

53. Rai V, Kumar P. Methylenetetrahydrofolate reductase C677T polymorphism and susceptibility to epilepsy. Neurol Sci 2018;39(12):2033–41.

54. Rai V. Evaluation of Methylenetetrahydrofolate Reductase Gene Variant (C677T) as Risk Factor for Bipolar Disorder. Cell Mol Biol 2011;57:1558–66.

55. Yadav U, Kumar P, Gupta S, Rai V. Role of MTHFR C677T gene polymorphism in the susceptibility of schizophrenia: An updated meta-analysis. Asian J Psychiatr 2016;20:41–51.

56. Rai V, Yadav U, Kumar P, Yadav SK, Gupta S. Methylenetetrahydrofolate reductase A1298C genetic variant&risk of schizophrenia: A meta-analysis. Indian J Med Res. 2017;145(4):437–47.

57. Rai V. Methylenetetrahydrofolate Reductase (MTHFR) C677T Polymorphism and Alzheimer Disease Risk: a Meta-Analysis. Mol Neurobiol 2017;54(2):1173–86.

58. Rai V. The methylenetetrahydrofolate reductase C677T polymorphism and breast cancer risk in Asian populations. Asian Pac J Cancer Prev 2014;15(14):5853–60.

59. Kumar P, Yadav U, Rai V. Methylenetetrahydrofolate reductase gene C677T polymorphism and breast cancer risk: Evidence for genetic susceptibility. Meta Gene 2015;6:72–84.

60. Rai V, Yadav U, Kumar P. Impact of Catechol-O-Methyltransferase Val 158Met (rs4680) Polymorphism on breast cancer Susceptibility in Asian population. Asian Pac J Cancer Prev 2017;18 (5):1243–50.

61. Rai V. Evaluation of the MTHFR C677T polymorphism as a risk factor for colorectal cancer in Asian populations. Asian Pac J Cancer Prev 2016;16(18):8093–100.

62. Yadav U, Kumar P, Rai V. “NQO1 Gene C609T Polymorphism (dbSNP: rs1800566) and Digestive Tract Cancer Risk: A Meta-Analysis.”. Nutr Cancer 2018;70(4):557–68.

63. Kumar P, Rai V. MTHFR C677T polymorphism and risk of esophageal cancer: an updated meta-analysis. Egypt J Med Hum Genet 2018;19:273–84.

64. Yadav U, Kumar P, Rai V. Role of MTHFR A1298C gene polymorphism in the etiology of prostate cancer: a systematic review and updated meta-analysis. Egypt J Med Hum Genet 2016;17(2):141–8.

